# Spatial Geometry and Prevalence of Tunneling and Undermining in Pressure Ulcers

**DOI:** 10.64898/2026.08.28.26361615

**Authors:** Sasha Frade, Zweli Tunyiswa, Michael Shin, Ryan Dirks

## Abstract

**Background:** Pressure ulcers often develop complex three-dimensional morphologies that extend beyond the visible wound surface. Subsurface extensions such as tunneling and undermining create hidden cavities that complicate clinical assessment and wound management. Despite their clinical relevance, the prevalence and spatial characteristics of these subsurface wound morphologies have not been well characterized at scale.

**Methods:** We performed a registry-based analysis using data from the LIFT-OFF Pressure Ulcer Registry, which captures longitudinal clinical documentation of pressure ulcers treated in routine care. The registry included approximately 18,000 patients with 32,000 documented pressure ulcers. Spatial characteristics of tunneling and undermining were analyzed using measurements recorded during routine wound assessments, including tract length, direction, and circumferential extent. Directional and circumferential distributions of subsurface defects were examined to characterize wound geometry.

**Results:** Tunneling was present in 764 of 14,700 full-thickness pressure ulcers (5.2%), whereas undermining occurred in 2,293 wounds (15.6%). Tunneling tracts were typically short and exhibited directional clustering relative to the wound bed. In contrast, undermining demonstrated broader circumferential distributions and frequently involved larger subsurface separations beneath the wound margin. Both morphologies demonstrated distinct spatial patterns across anatomical locations and wound stages.

**Conclusion:** Tunneling and undermining are common subsurface features of pressure ulcers and exhibit distinct spatial geometries. Whereas tunneling manifests as directional tract-like extensions, undermining more frequently produces circumferential tissue separation beneath wound margins. Improved characterization of subsurface wound architecture may enhance assessment of wound complexity and provide information not captured by surface measurements alone. Future studies should evaluate whether these features contribute to wound severity assessment, prognosis, and risk stratification.

## I. Introduction

Pressure ulcers frequently develop complex three-dimensional morphologies that extend beyond the visible wound surface.^1–3^ In addition to surface tissue loss, wounds may develop subsurface extensions that alter wound architecture and complicate both assessment and management. Two commonly observed manifestations of such subsurface extension are tunneling and undermining. Tunneling refers to a narrow tract that extends from the wound base into adjacent tissue, whereas undermining reflects separation of tissue beneath the wound edge, creating a cavity that extends laterally under intact skin margins.^1,4^ Both phenomena create subsurface cavities beneath the visible wound surface that are not captured by simple surface measurements. A schematic representation of these subsurface wound cavities is illustrated in Figure 1.

**Figure 1.**
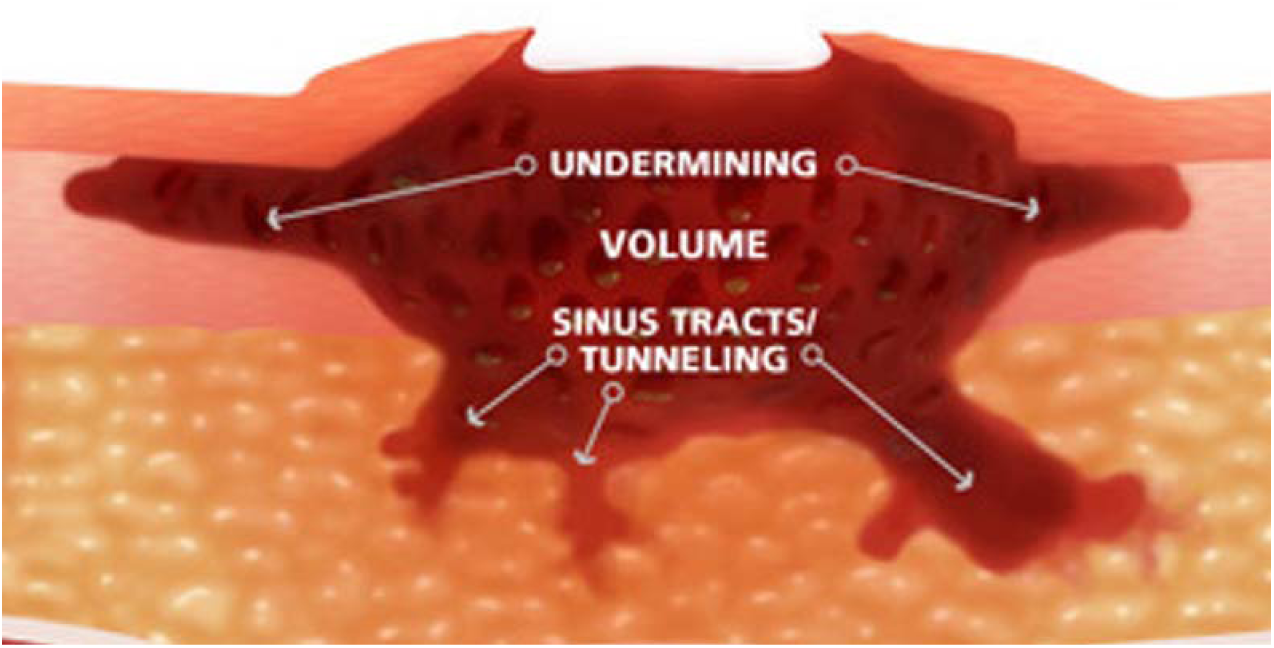
Schematic representation of tunneling and undermining in pressure ulcers.

Although tunneling and undermining are often grouped together in clinical descriptions of wound morphology, they represent distinct geometric forms of tissue loss. Tunneling can be conceptualized as a directed tract characterized by both magnitude and direction, analogous to a vector extending from the wound base into surrounding tissue. In contrast, undermining represents circumferential separation of tissue beneath the wound edge and can be conceptualized as an arc extending laterally around the wound perimeter. This distinction between a directional tract and a circumferential extension provides a useful geometric framework for describing subsurface wound morphology and may help clarify how these defects develop and propagate within tissue. Recognizing these geometric differences may be important for understanding how subsurface defects influence wound structure and behavior within the tissue environment. For spatial analysis, tunneling was conceptualized as a directional vector defined by magnitude and orientation, whereas undermining was represented as a circumferential arc defined by angular extent around the wound margin (Box 1).

**Box 1.**
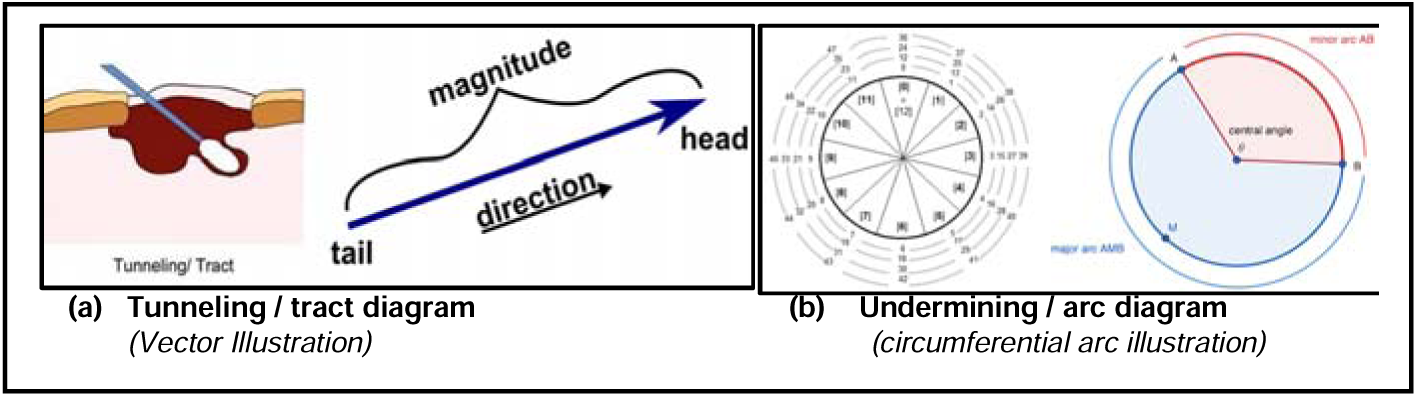
Conceptual geometric framework used for spatial characterization of subsurface wound defects. (a) Tunneling is represented as a directional tract extending from the wound base and can be described by both magnitude (tract length) and direction relative to the wound bed. (b) Undermining represents circumferential tissue separation beneath the wound margin and is

The presence of tunneling and undermining introduces subsurface cavities that complicate wound management. These hidden spaces may impair effective contact between dressings and the wound bed, potentially limiting the ability of topical therapies to interact with the affected tissue. In addition, subsurface cavities may contribute to persistent dead space within the wound environment, which can interfere with granulation and tissue repair.^5,6^ Because many wound care approaches are designed primarily to cover the wound surface, they may not fully address the structural voids created by these subsurface extensions. Therapies capable of conforming to or occupying these subsurface spaces may therefore have potential to improve restoration of wound architecture and support tissue repair.

Despite their clinical relevance, the epidemiology and spatial characteristics of tunneling and undermining remain incompletely described. Chronic wounds represent a substantial and growing healthcare burden worldwide.^7,8^ However, most existing reports focus on individual cases or small cohorts and provide limited information regarding how frequently these morphologies occur across large populations of pressure ulcers.^1,4^

Similarly, relatively little is known about their spatial characteristics, including tract length, circumferential extent, and directional distribution. A more systematic characterization of these features could improve understanding of wound morphology and inform clinical management.

The objective of this study was to characterize the prevalence and spatial geometry of tunneling and undermining in pressure ulcers using data from a large clinical wound registry. Improved understanding of these spatial features may provide a foundation for future studies examining their clinical implications.

## II. Methods

### Data source and study population

Data for this analysis were derived from the LIFT-OFF Pressure Ulcer Registry, a large clinical wound registry capturing longitudinal data from patients receiving treatment for pressure ulcers in routine clinical practice. The registry includes structured documentation of wound characteristics recorded during routine clinical assessments, including wound size, stage, anatomical location, and the presence of subsurface extensions such as tunneling and undermining. The registry contains de-identified clinical data collected as part of routine wound care documentation. Because the present analysis used de-identified real-world data and did not involve direct patient interaction, institutional review board approval was not required.

At the time of analysis, the registry contained records for approximately 18,000 patients and 32,000 pressure ulcers. The registry prospectively collects standardized wound assessment data across participating clinical centers. These registry assessments captured standard wound characteristics as well as measurements related to subsurface wound morphology.

The study population consisted of pressure ulcers documented within the registry with recorded wound assessments. For descriptive analyses of subsurface wound morphology, all pressure ulcers with documented evaluation of tunneling or undermining were eligible for inclusion.

A subset analysis was performed for full-thickness pressure ulcers, including Stage 3, Stage 4, and unstageable pressure ulcers demonstrating full-thickness tissue loss. This subset included approximately 10,000 patients and 14,700 wounds. Patient demographics, wound stage, and anatomical location were summarized descriptively where available.

### Definitions, measurements, and spatial characterization

Tunneling was defined as a narrow tract extending from the wound base into adjacent tissue. When present, tunneling was documented with measurements of tract length and direction relative to the wound bed. Within the registry, tunneling direction was recorded using the standard clinical clockface convention relative to the wound bed, and undermining was documented using the recorded circumferential extent around the wound margin.

Undermining was defined as separation of tissue beneath the wound edge resulting in a cavity extending laterally beneath intact skin margins. Undermining was documented using circumferential measurements describing the extent of tissue separation around the wound perimeter.

Tunneling and undermining measurements were recorded as part of routine wound assessments within the registry. The present spatial analyses were performed using these documented measurements. Directional and circumferential representations used in this study were analytical constructs developed from available registry variables to characterize subsurface wound geometry and were not recorded directly as independent clinical variables within the registry. Recorded directional and circumferential measurements were standardized for analysis to facilitate pooled comparisons across wounds and anatomical locations. As with all registry-based wound assessments, measurements may be subject to interobserver variability and differences in documentation practices across clinicians and participating sites.

Distances associated with tunneling and undermining were recorded during routine wound assessments and used to characterize the spatial extent of these subsurface defects. Tunneling was analyzed as a directional structure defined by both tract length and orientation relative to the wound bed. Undermining was characterized by its circumferential extent around the wound margin, reflecting the arc-like distribution of tissue separation beneath the wound edge.

These measurements were used to examine the distribution and spatial patterns of subsurface wound defects across the study population.

### Analysis methods

Descriptive analyses were first performed to characterize the prevalence of tunneling and undermining among pressure ulcers recorded in the registry. The proportion of wounds exhibiting tunneling and undermining was calculated for the overall study population.

The spatial extent of these subsurface defects was then evaluated. Distributions of tunneling tract length and undermining extent were summarized to characterize the magnitude of subsurface tissue loss. Directional patterns of tunneling were subsequently examined based on recorded tract orientation relative to the wound bed. Circumferential patterns of undermining were analyzed based on the documented arc of tissue separation around the wound margin.

The analyses were descriptive and exploratory in nature and were intended to characterize the prevalence and spatial patterns of subsurface wound morphology. No formal inferential statistical comparisons were prespecified across anatomical locations, wound stages, or spatial distributions. Accordingly, observed differences should be interpreted as descriptive and hypothesis-generating.

## III. Results

Characteristics of the study population are summarized in Table 1. The registry included approximately 18,000 patients with 32,000 documented pressure ulcers recorded during routine clinical care. Among these, a subset of approximately 10,000 patients with 14,700 full-thickness pressure ulcers was included in the spatial morphology analyses.

Subsurface extensions were commonly observed among pressure ulcers recorded in the registry. Among full-thickness pressure ulcers, tunneling was documented in 764 of 14,700 wounds (5.2%), whereas undermining was documented in 2,293 of 14,700 wounds (15.6%). Thus, undermining was observed approximately three times more frequently than tunneling within the registry population.

As shown in Figure 2, the prevalence of both tunneling and undermining varied by anatomical location. Tunneling occurred most frequently in pressure ulcers located at the hip (12.7%) and back (7.4%), with lower rates observed in ulcers of the buttock (3.9%), ankle (1.7%), and heel (0.5%). A similar anatomical pattern was observed for undermining, although at substantially higher rates. Undermining was most frequently observed in ulcers located at the hip (30.4%) and back (24.6%), followed by the buttock (10.7%) and elbow (8.9%), with lower rates in ulcers of the ankle (3.7%) and heel (2.4%).

**Figure 2.**
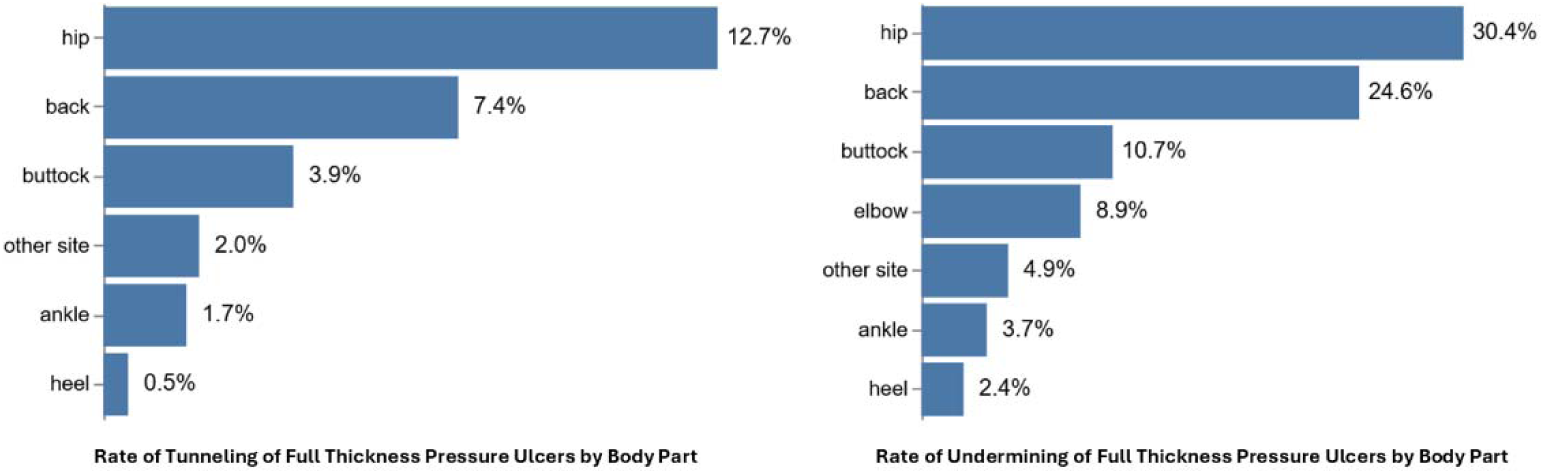
Prevalence of tunneling and undermining among full-thickness pressure ulcers by anatomical location.

Across nearly all anatomical locations, undermining occurred more frequently than tunneling, suggesting that circumferential tissue separation beneath the wound margin represents a more common form of subsurface tissue loss than tract-like extensions from the wound base.

The spatial extent of subsurface defects differed between tunneling and undermining. Figure 3 shows that tunneling tracts were generally short, with the majority of tunneling measurements occurring within the first 1–2 cm of extension from the wound base. The cumulative distribution indicates that most tunneling tracts were relatively limited in length, with only a small proportion extending beyond several centimeters.

**Figure 3.**
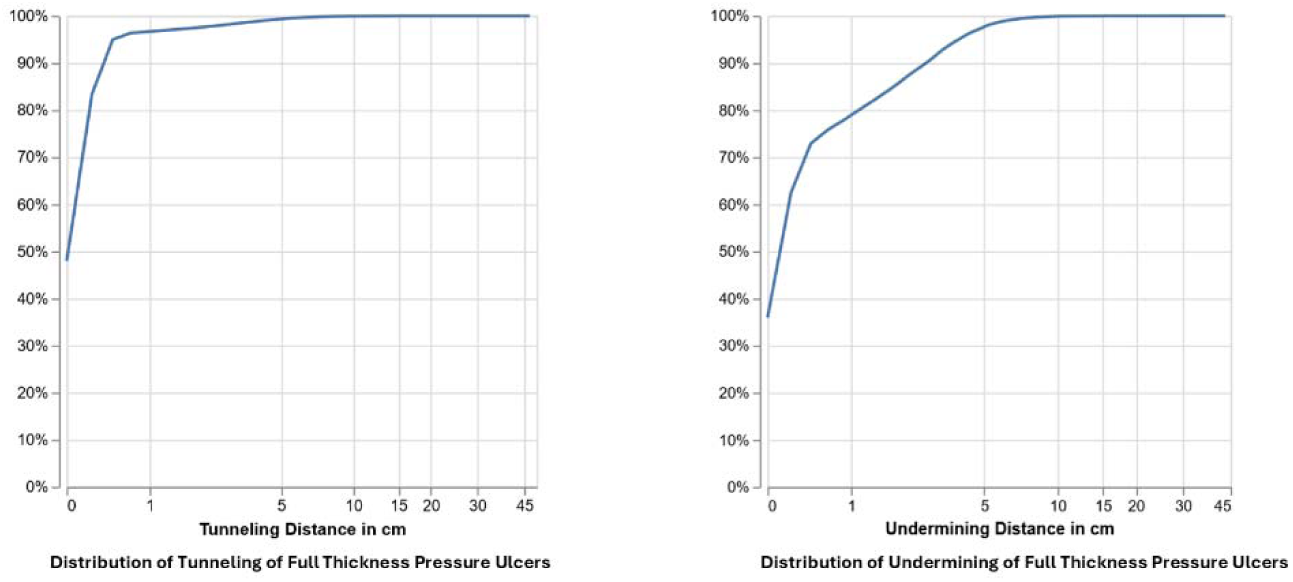
Cumulative distribution of tunneling tract length and undermining extent among full-thickness pressure ulcers.

In contrast, undermining demonstrated a broader distribution of spatial extent. Although a substantial proportion of undermining measurements occurred within the first few centimeters, the cumulative distribution increased more gradually compared with tunneling, indicating that larger subsurface separations beneath the wound margin were more frequently observed. This pattern suggests that undermining not only occurs more frequently than tunneling but also tends to involve greater lateral tissue separation.

Together, these findings highlight the distinct spatial characteristics of these two subsurface morphologies: tunneling typically manifests as short, tract-like extensions, whereas undermining more commonly produces broader circumferential cavities beneath intact wound margins.

Directional analyses of tunneling demonstrated identifiable orientation patterns relative to the wound bed. Figure 4 demonstrates that tunneling tracts were not uniformly distributed across directions. Instead, a substantial proportion of observations were concentrated within superior clock-face positions relative to the wound bed, indicating a dominant directional pattern of tract propagation.

**Figure 4.**
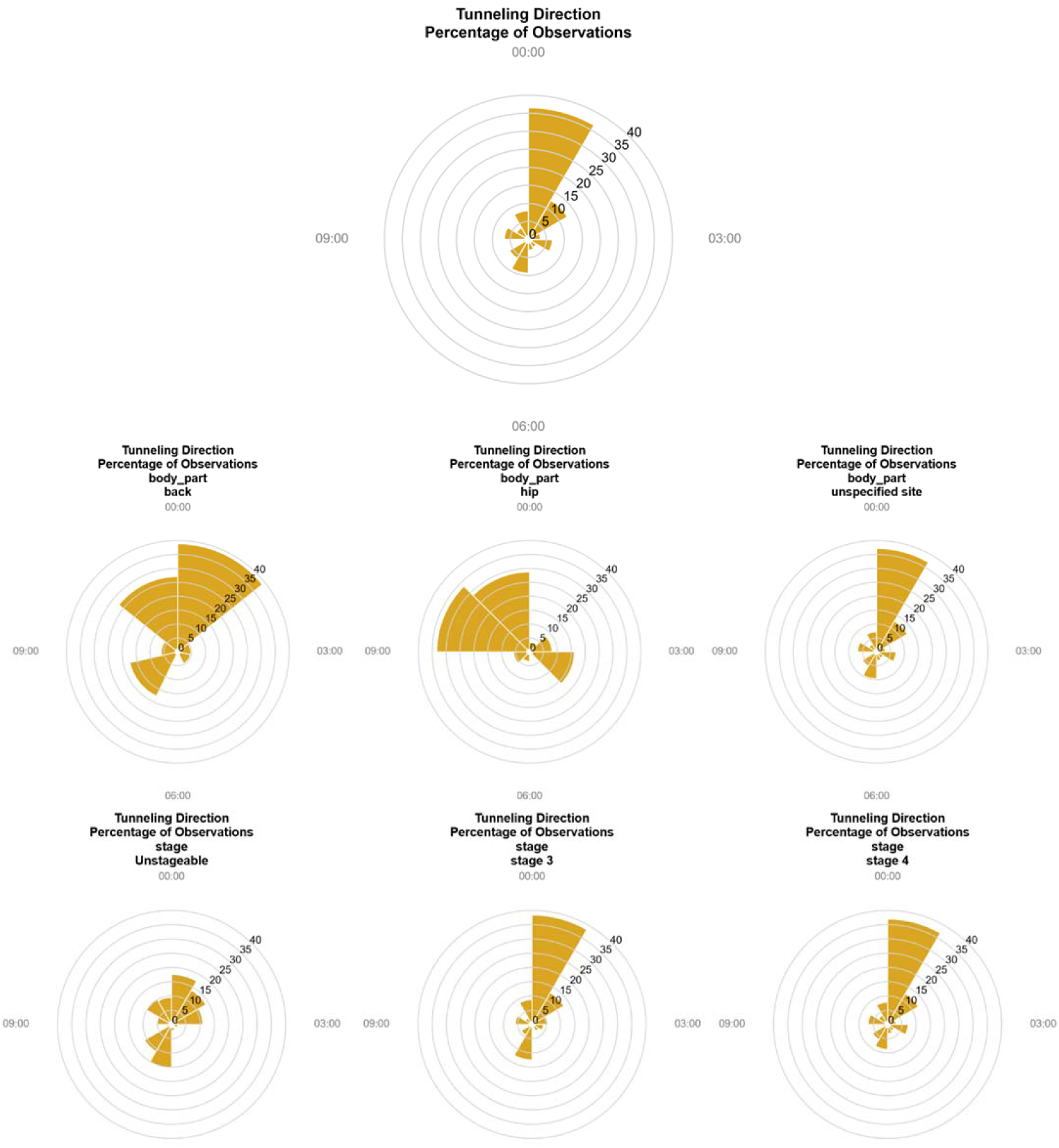
Directional distribution of tunneling tracts relative to the wound bed, shown for the overall dataset and stratified by anatomical location and wound stage.

This directional clustering was observed in the overall dataset and remained evident when tunneling direction was examined across anatomical locations and wound stages. Similar directional patterns were observed for wounds located at the back and hip, which together accounted for a large proportion of tunneling observations. Comparable orientation patterns were also evident across Stage 3 and Stage 4 pressure ulcers, suggesting that tunneling tends to propagate along consistent tissue planes regardless of wound severity.

These findings support the characterization of tunneling as a directional tract-like extension, consistent with the conceptualization of tunneling as a vector defined by both magnitude and orientation.

Circumferential distributions of undermining were analyzed based on the documented arc of tissue separation around the wound perimeter. As shown in Figure 5, undermining was observed across all circumferential positions, indicating that lateral tissue separation beneath the wound margin can occur around the entire wound perimeter. However, the distribution was not uniform.

**Figure 5.**
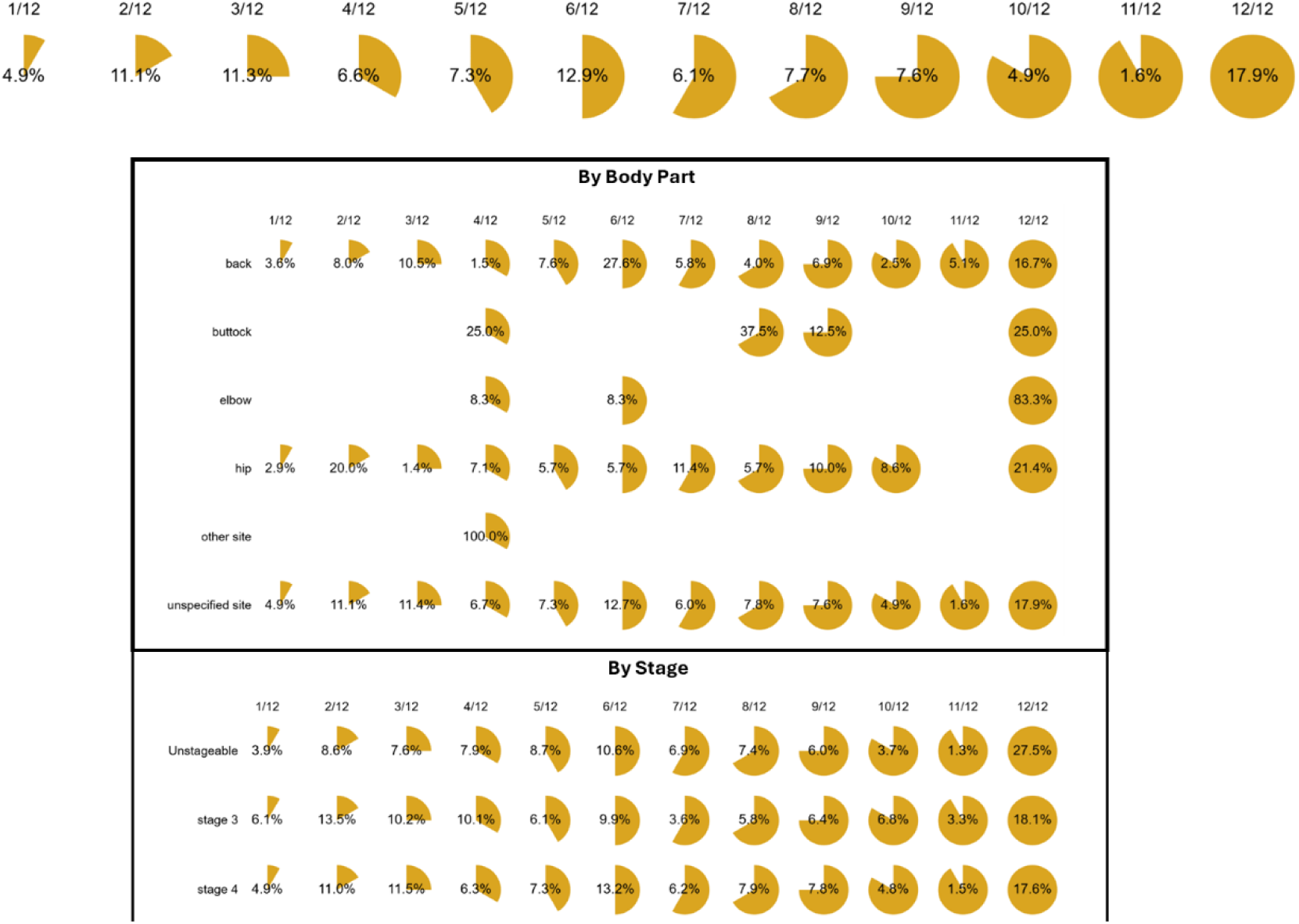
Circumferential distribution of undermining around the wound margin, shown overall and stratified by anatomical location and wound stage.

Across the overall dataset, certain circumferential segments demonstrated higher proportions of undermining observations than others, indicating that undermining tends to concentrate within particular arcs of the wound margin. When stratified by anatomical location, similar but site-specific patterns were observed. For example, wounds located at the back and hip demonstrated broader distributions of undermining across multiple circumferential segments, whereas other sites showed more localized patterns of tissue separation.

Comparable circumferential distributions were observed across wound stages. Both Stage 3 and Stage 4 pressure ulcers demonstrated undermining across multiple positions around the wound margin, suggesting that circumferential tissue separation represents a common structural feature of advanced pressure ulcers.

These findings contrast with the directional tract-like patterns observed for tunneling and support the characterization of undermining as a circumferential extension of tissue separation beneath the wound edge.

## IV. Discussion

In this registry-based analysis of pressure ulcers, tunneling and undermining were common manifestations of subsurface wound morphology but demonstrated distinct patterns of prevalence and spatial geometry. Undermining occurred approximately three times more frequently than tunneling within the registry population, indicating that lateral tissue separation beneath the wound margin represents a common structural feature of pressure ulcers. In addition to differences in prevalence, these morphologies exhibited distinct spatial characteristics. Tunneling appeared as directional tract-like extensions from the wound base into adjacent tissue, whereas undermining manifested as circumferential separation beneath intact wound margins. Together, these findings support the concept that subsurface wound defects represent structured geometric features rather than purely descriptive clinical observations.^1–4^

The spatial patterns identified in this analysis suggest that tunneling and undermining represent distinct modes of subsurface tissue disruption. Tunneling behaves as a tract-like extension that propagates directionally from the wound base, whereas undermining reflects circumferential tissue separation beneath intact skin margins. These structural differences may reflect underlying biomechanical forces, pressure distribution, or local tissue perfusion that contribute to the progression of pressure-related tissue injury.^1–3, 9^ Recognizing these geometric distinctions may therefore provide a more structured framework for describing subsurface wound morphology in clinical assessment and research.

The presence of tunneling and undermining introduces subsurface cavities that may complicate wound management. Because these spaces extend beyond the visible wound surface, they may not be fully captured by conventional surface measurements used to document wound size or progression. In addition, subsurface cavities may impair effective contact between topical therapies and the wound bed. Persistent dead space within these cavities can interfere with granulation tissue formation and wound contraction, both of which are essential components of wound healing.^5,6,10^ As a result, the ability to properly assess and measure subsurface wound morphology may represent an underrecognized factor in the evaluation and management of pressure ulcers.

Beyond their implications for wound management, tunneling and undermining may represent clinically meaningful dimensions of wound complexity that are not adequately captured by conventional surface measurements alone. Current pressure injury assessment frameworks rely heavily on visible wound characteristics, tissue involvement, and stage classification to evaluate severity and guide management.^1,14^ However, wounds with extensive subsurface extension may contain substantially greater tissue destruction than is apparent from surface dimensions, resulting in larger volumes of tissue loss, increased dead space, and more complex wound architecture.^1–6^ As a consequence, wounds with similar surface dimensions and stage classifications may differ considerably in biological burden and healing potential if one contains extensive tunneling or undermining while the other does not.

This observation raises the possibility that subsurface wound morphology may provide prognostic information beyond traditional wound measurements. Previous studies have demonstrated that wound size, depth, tissue loss, and early healing trajectories are important predictors of healing outcomes in chronic wounds.^12,13,15^ Whether tunneling length, undermining extent, or specific spatial configurations independently contribute to prognosis remains unknown. Future studies linking subsurface wound architecture to healing, recurrence, infection, hospitalization, and other clinically important outcomes may help determine whether these features represent independent markers of wound severity and complexity. If confirmed, incorporation of subsurface wound characteristics into wound assessment frameworks could improve severity classification, risk stratification, prognosis, and treatment planning.

The geometric characteristics identified in this study highlight potential limitations of treatment approaches that primarily address the visible wound surface. Conventional dressings are typically designed to cover the wound bed but may not fully conform to or occupy subsurface cavities created by tunneling or undermining. Therapeutic materials capable of conforming to complex wound geometries or filling subsurface voids may therefore provide advantages in wounds characterized by these features. Directional clustering of tunneling tracts observed in this analysis further supports the concept that tunneling propagates along preferential tissue planes rather than occurring randomly within the wound bed. Improved characterization of subsurface wound geometry may help inform the development of treatment approaches that better address the three-dimensional structure of chronic wounds.^5–8,11^

This study has several strengths. The analysis was performed using data from the LIFT-OFF Pressure Ulcer Registry, which contains longitudinal documentation of a large number of pressure ulcers recorded during routine clinical care. This large dataset enabled characterization of subsurface wound morphology across a broad population of wounds. An additional implication of these findings is that subsurface wound architecture may represent a previously underutilized source of information within wound registries and electronic health records. Most contemporary wound datasets emphasize surface dimensions and tissue characteristics, whereas tunneling and undermining are often documented descriptively and infrequently incorporated into prognostic models. The present findings suggest that these features may warrant further investigation as candidate variables in future predictive models of wound healing and treatment response.

However, several limitations should also be acknowledged. Registry-based analyses rely on clinical documentation recorded during routine care, and measurement techniques for tunneling and undermining may vary among clinicians. In addition, the directional and circumferential spatial representations used in this analysis were derived from routine clinical documentation rather than collected as dedicated research measurements. Consequently, variability in wound assessment and documentation practices may have influenced characterization of subsurface wound geometry. Furthermore, the observational nature of registry data limits causal inference regarding the role of subsurface wound morphology in pressure ulcer progression and clinical management. Future prospective studies may further clarify how these geometric features influence wound progression and response to therapy.

Future studies should evaluate how these geometric characteristics relate to wound healing outcomes and treatment response. Understanding the relationship between subsurface wound architecture and healing dynamics may further inform the development of targeted therapeutic strategies.

## V. Conclusions

In this large registry-based analysis, tunneling and undermining were common features of pressure ulcers and demonstrated distinct spatial geometries. Whereas tunneling typically manifested as directional tract-like extensions from the wound base, undermining more frequently involved broader circumferential tissue separation beneath the wound margin. These findings highlight the importance of characterizing subsurface wound architecture when evaluating complex wounds. Improved assessment of tunneling and undermining may enhance understanding of wound complexity and provide information not fully captured by conventional surface measurements alone. Future studies should evaluate whether these subsurface morphological features are associated with wound healing outcomes and whether they contribute independently to severity assessment, prognosis, and risk stratification. Improved characterization of subsurface wound architecture may help inform clinical management and guide the development of therapeutic strategies designed to address the three-dimensional morphology of chronic wounds.

## Author Contributions

Conceptualization, ZT; methodology, ZT; formal analysis, ZT; data curation, ZT; writing—original draft preparation, SF; writing—review and editing, all authors. All authors have read and agreed to the published version of the manuscript.

## Institutional Review Board Statement

Ethical review and approval were waived for this study because the analysis used de-identified real-world clinical registry data collected as part of routine clinical documentation and did not involve direct patient interaction or identifiable patient information.

## Informed Consent Statement

Patient consent was waived because the study used de-identified registry data collected during routine clinical care and did not involve identifiable patient information.

## Data Availability Statement

The data analyzed in this study were obtained from the LIFT-OFF Pressure Ulcer Registry. Access to the data is subject to registry governance and data use agreements and is therefore not publicly available. Data may be available from the corresponding author upon reasonable request and with permission of the registry data custodians.

## Acknowledgments

The authors acknowledge the clinicians and participating centers contributing data to the LIFT-OFF Pressure Ulcer Registry, whose routine clinical documentation made this analysis possible.

